# ASSESSMENT OF PERFORMANCE AND IMPLEMENTATION CHARACTERISTICS OF RAPID POINT OF CARE SARS-CoV-2 ANTIGEN TESTING

**DOI:** 10.1101/2021.06.03.21258290

**Authors:** Eva Muthamia, Samuel Mungai, Mary Mungai, Gama Bandawe, Firdausi Qadri, Zannat Kawser, Shahin Lockman, Louise C. Ivers, David Walt, Sara Suliman, Matilu Mwau, Jesse Gitaka

## Abstract

**Background:** The COVID-19 pandemic has resulted in a need for rapid identification of infectious cases. Testing barriers have prohibited adequate screening for SARS COV2, resulting in significant delays in treatment provision and commencement of outbreak control measures. This study aimed to generate evidence on the performance and implementation characteristics of the BD Veritor rapid antigen test as compared to the gold standard test for diagnosis of SARS COV2 in Kenya.

**Methods:** This was a field test performance evaluation in symptomatic and asymptomatic adults undergoing testing for SARS COV2. Recruited participants were classified as SARS-CoV2-positive based on the locally implemented gold standard reverse transcription polymerase chain reaction (RT-PCR) test performed on nasopharyngeal swabs. 272 antigen tests were performed with simultaneous gold standard testing, allowing us to estimate sensitivity, specificity, positive and negative predictive values for the BD Veritor rapid antigen test platform. Implementation characteristics were assessed using the Consolidated Framework for Implementation Research for feasibility, acceptability, turn-around time, and ease-of-use metrics.

**Results and Discussion:** We enrolled 97 PCR negative symptomatic and 128 PCR negative asymptomatic, and 28 PCR positive symptomatic and 19 PCR positive asymptomatic participants. Compared to the gold standard, the sensitivity of the BD Veritor antigen test was 94% (95% confidence interval [CI] 86.6 to 100.0) while the specificity was 98% (95% confidence interval [CI] 96 to 100). The sensitivity of BD Veritor antigen test was higher among symptomatic (100%) compared to asymptomatic (84%) participants, although this difference was not statistically significant. There was also a lack of association between cycle threshold value and sensitivity of BD Veritor test. The BD Veritor test had quick turnaround time and minimal resource requirements, and laboratory personnel conducting testing felt that it was easier to use than the gold standard RT-PCR.

**Conclusion:** The BD Veritor rapid antigen test exhibited excellent sensitivity and specificity when used to detect SARS-CoV-2 infection among both symptomatic and asymptomatic individuals in varied population settings in Kenya. It was feasible to implement and easy to use, with rapid turnaround time.

## INTRODUCTION

The coronavirus disease of 2019 (COVID-19) has placed enormous burdens on individuals and society at large. Low and middle-income countries in particular are disadvantaged as resources are already significantly stretched. Huge surges in infection carry the risk of quickly overwhelming health care systems, leading to excess mortality. Therefore, rapid identification and isolation of infectious cases is key to containing the pandemic. Real-time reverse transcription polymerase chain reaction (RT-PCR) has been the reference standard method for detection of Severe Acute Respiratory Syndrome Coronavirus-2 (SARS-CoV-2) infection. Despite its excellent sensitivity and specificity, the method is limited by high initial set up costs, expensive consumables, need for highly trained staff, prolonged turn-around time, requirement for sample and results transport and an uninterrupted power supply(1). Recent evidence indicates test reporting delays as having a negative impact on isolation as a control measure of infection spread (2). There is therefore need to optimize testing modalities that can be applied to large populations quickly enough to inform strategies that limit transmission(3)(4).

The exigency for decentralized testing options and rapid development of novel biomarkers has resulted in the development and approval of rapid antigen tests as a complementary modality to RT-PCR (5). They are less costly, can easily be offered at the point of care, have fewer associated health worker training needs, and likely identify the most infectious individuals early in disease course. The use of rapid antigen tests can improve accessibility to testing, facilitate timely confirmation of suspected cases and expedite clinical and public health decision making.

However, rapid antigen test performance varies depending on inherent test characteristics, quality of sample, timing of sample collection in disease course, SARS-CoV-2 viral load and presence of symptoms (6).

While viral RNA can be detected by RT-PCR weeks after infection, culture-positive specimens are generally not found after 9 days post-infection. Culture-positive samples contain more viral RNA than culture negative specimens (7)(8). Peak RNA concentrations are reached before day 5 of symptom onset, and the potential for transmission declines after 1 week of symptom onset(9)(8). RT-PCR amplifies and detects nucleic acids, including sub-genomic RNA that represent non-intact virus (10). As cycle threshold (Ct) value and viral load levels are inversely correlated, samples with high Ct values on RT-PCR are from individuals who are less likely to be contagious (9).

Sensitivity of rapid antigen tests appears to generally be higher in symptomatic patients and in those with high viral loads (6) (11) (12). These are patients who are more likely to be infectious. Rapid antigen test sensitivity is also higher when carried out less than 5 days from symptom onset. The overall low sensitivity of rapid antigen tests studies has been attributed to false negative results seen in samples with high RT-PCR Ct values (13). There is a low probability of transmission from patients whose samples test positive on RT-PCR but negative on rapid antigen tests. (14). The World Health Organization (WHO) recommends that rapid antigen tests should have a sensitivity of >/=80% and a specificity of >/=97% (15). At a population level, the lower sensitivity of rapid tests may be improved by high frequency testing(16)(17).

It is necessary to evaluate rapid antigen tests at the end-user level, taking the local population into consideration before large-scale implementation (18). Alignment with existing health systems is a determinant for adoption of novel diagnostic methods. Besides technical performance, evaluation of rapid antigen tests should include aspects related to clinical utility, cost and patient satisfaction (19). This information is useful to key stakeholders such as researchers, product developers, payers and policy makers in understanding the real-world context so as to meet the needs of COVID-19 testing.

The BD Veritor System for Rapid Detection of SARS-CoV-2 is a rapid chromatographic immunoassay for the detection of SARS-CoV-2 antigens in respiratory specimens. Viral nucleocapsid protein is targeted for detection(20). This study aims to evaluate the performance and implementation characteristics of BD Veritor rapid antigen test compared to gold-standard RT-PCR in asymptomatic and symptomatic adults undergoing testing for SARS-CoV-2 in Kenya. Findings from this study will inform the design of SARS CoV2 testing protocols and guide large scale use of rapid antigen tests.

## METHODS

### Study design and participants

Individuals aged 18 and above who were being tested for SARS-CoV-2 in Kenya as part of their routine care, and who gave written consent for participating in this study were enrolled consecutively between 31^st^ January and 24^th^ March 2021. The participants included travelers, university students, healthcare workers (HCWs), patients seeking services in hospital outpatient departments (OPD) and members of the general population. Healthcare workers and OPD patients were enrolled at Mary Help Hospital in Thika, Kenya, while students and general population were enrolled at Mount Kenya University in Thika, Kenya. Our enrollment targets were 100 PCR negative symptomatic and asymptomatic participants each, and 30 PCR positive symptomatic and asymptomatic participants each. Known SARS-CoV-2 RT-PCR positive participants were retrospectively identified from laboratory records and invited to consent for re-testing within 24 hours of collection of the initial RT-PCR positive sample. These were mainly individuals that required SARS-CoV-2 testing before travel. At re-test, samples for RT-PCR and rapid antigen test were collected. Both symptomatic and asymptomatic PCR positive and negative participants were enrolled, and recruitment was carried out irrespective of duration of symptoms. Demographic and clinical information was obtained using clinical evaluation and an interviewer-administered questionnaire.

Ethical approval for this study was granted by the Mount Kenya University Ethics Review Committee (MKU/ERC/1780).

### Sample collection

Paired oropharyngeal and anterior nasal swabs were obtained in the same encounter for RT-PCR and rapid antigen testing respectively. For the rapid antigen test, anterior nasal specimens were obtained using regular-tipped flocked swabs inserted approximately 2-3cm into the anterior nares. The swab was rolled along the mucosa of each nostril. The specimen obtained was then processed according to the manufacturer’s recommendations.

Samples for RT-PCR were obtained via oropharyngeal swabs. They were placed in viral transport medium and delivered in cooler boxes at 2 to 8 Degrees Celsius to the Kenya Medical Research Institute (KEMRI) laboratory. Prior to testing, samples were removed from the cooler boxes and allowed to reach room temperature. Samples from asymptomatic participants were analyzed in pooled samples of 10, while those from symptomatic participants were run singly. This was in accordance with standard operating procedures at the laboratory. The RT-PCR assay used was

Abbott RealTime SARS-CoV-2 (21). The target sequences were the SARS-CoV-2 RNA-dependent RNA polymerase (RdRp) and N genes. A positive result was confirmed when either a single-gene or a two-gene amplification occurred. Positive and negative internal control were included in each run. Test results were interpreted as positive or negative at a Ct of 37 according to the manufacturer’s recommendation.

### Qualitative data collection

Trained research assistants collected qualitative data using face-to-face key-informant interviews. Semi-structured interview guides were used. Key informant interviews were conducted in three population subsets; travelers, individuals seeking care in hospital out-patient departments and health care workers offering testing services. Seven Key Informant Interviews (KIIs) with health care workers, seven KIIs with hospital clients and three KIIs with travelers were conducted. All three population subsets were interviewed on their perceptions on barriers and facilitators, satisfaction, ease of use and acceptability of BD Veritor antigen test.

### Statistical analysis

The primary pre-specified outcome measures for this study were sensitivity and specificity point estimates and 95% confidence intervals for the BD Veritor antigen test compared to results from the reference standard oropharyngeal swab RT-PCR. Statistical analysis was performed using SPSS version 23.0 software. Overall sensitivity and specificity of BD Veritor antigen test was calculated and then stratified between asymptomatic and symptomatic individuals. The diagnostic measures efficiency of the BD Veritor antigen test was further correlated with the Ct threshold values of RT-PCR and sensitivity stratified by persons with low and high Ct values. 95% confidence intervals (CI) were calculated for all the sensitivity and specificity proportions. Participant’s characteristics were summarized and presented as percentages. The differences in participants’ characteristics based on PCR positivity status and presence of symptoms was explored and tested using chi square test of associations. Age of the participants was presented as mean and compared between groups using Student’s t test. P value less or equal to 0.05 was statistically significant.

Qualitative data was captured using audio tapes and field notes, transcribed and managed using QSR NVivo 12 software. The KII transcripts were coded and checked for coding consistency using a thematic framework to classify and organize data into four themes. These included knowledge on COVID-19 testing, sample collection experience, applicability of the rapid antigen test and improvement suggestions for the test. We applied a grounded theory approach (22). We used an iterative process to develop the thematic framework and updated in two rounds of analysis. Analysis charts for each emergent theme were developed and categorized across all participants.

## RESULTS

287 participants were enrolled into the study. 15 did not meet the eligibility criteria for age and were not included in the analysis. 272 paired samples were obtained. The participants had a median age of 30 years (range 18 to 68), 135 (50%) were female and 125 (46%) participants were symptomatic (by design). 47 (17%) samples tested positive on RT-PCR while 49 (18%) were positive on BD Veritor antigen test. Health Care Workers (HCWs) and Out-Patient Department (OPD) patients comprised the majority of symptomatic participants (33% each), while students comprised majority of asymptomatic participants (54%). The median duration of symptoms was 5 days **(Table 1)**.

**Table 1:** Participants’ characteristics.

| **Variable** | **Frequency (%)** | **PCR negative** | **PCR positive** | **P value** |
| --- | --- | --- | --- | --- |
| **Sex** |  |  |  |  |
| Female | 135 (50) | 114 (51) | 21 (45) | 0.455 |
| Male | 137 (50) | 111 (49) | 26 (55) |  |
| Mean age in years (SD) | 30 (10) | 29 (9) | 37 (12) | <0.001 |
| Min-Max | 18-68 |  |  |  |
| **Population** |  |  |  |  |
| General | 30 (11) | 16 (7) | 14 (30) |  |
| HCW | 75 (28) | 60 (27) | 15 (32) |  |
| OPD patient | 41 (15) | 41 (18) | 0 |  |
| Student | 90 (33) | 89 (40) | 1 (2) |  |
| Traveler | 36 (13) | 19 (8) | 17 (36) |  |
| **Clinical status** |  |  |  |  |
| Asymptomatic | 147 (54) | 128 (57) | 19 (40) | 0.039 |
| Symptomatic | 125 (46) | 97 (43) | 28 (60) |  |
| **Number of symptoms** |  |  |  |  |
| 0 | 147 (54) | 128 (57) | 19 (40) | 0.021 |
| 2 | 2 (1) | 1 (0) | 1 (2) |  |
| 3 | 1 (0) | 0 | 1 (2) |  |
| More | 122 (50) | 96 (43) | 26 (55) |  |
| **Duration of symptoms in days (n=76)** |  |  |  |  |
| Median (IQR) | 5 (2-7) | 4 (1.5-6.5) | 5 (2.5-7.5) | 0.399 |
| **Category, n (%)** |  |  |  |  |
| 5 days or less | 42 (55) | 32 (57) | 10 (50) | 0.581 |
| More than 5 days | 34 (45) | 24 (43) | 10 (50) |  |
| **Type of symptoms** |  |  |  |  |
| Sore throat | 53 (60) |  |  |  |
| Headache | 48 (55) |  |  |  |
| Cough | 46 (52) |  |  |  |
| Runny nose | 39 (44) |  |  |  |
| General Weakness | 33 (38) |  |  |  |
| Fever/Chills | 32 (36) |  |  |  |
| Shortness of breath | 18 (21) |  |  |  |
| Muscular pain | 17 (20) |  |  |  |
| Nausea/Vomiting | 14 (16) |  |  |  |
| Joint pain | 13 (15) |  |  |  |
| Abdominal pain | 12 (14) |  |  |  |
| Chest pain | 11 (13) |  |  |  |
| Diarrhoea | 9 (10) |  |  |  |
| Irritability/Confusion | 7 (8) |  |  |  |

Abbreviations: PCR, Polymerase Chain Reaction; HCW, Health Care Worker; OPD, Out-Patient Department; SD, Standard Deviation; IQR, Inter Quartile Range

Compared to RT-PCR, the sensitivity of the BD Veritor antigen test was 94% (95% confidence interval [CI] 87 to 100) while the specificity was 98% (95% confidence interval [CI] 96 to 100). Overall concordance was 97% (95% confidence interval [CI] 95 to 99) from 264/272 specimens **(Table 2)**. The sensitivity of BD Veritor antigen test was higher among symptomatic compared to asymptomatic participants (100% vs. 84%), although this did not reach statistical significance. Likewise, no significant difference in specificity was observed between symptomatic (96%) and asymptomatic (99%) participants **(Table 3)**. There was no statistical difference in qualitative PCR results (p=0.581), or quantitative Ct values (p=0.840) and sensitivity of the rapid antigen test between those who had symptoms for less than 5 days (inclusive) and those who had symptoms for more than 5 days.

**Table 2:** Performance of BD Veritor antigen test against RT-PCR.

| Results |  | BD performance | 95% CI |
| --- | --- | --- | --- |
| Positive | PCR | 47 | - |
|  | BD | 44 | - |
|  | Sensitivity (PPA) | 94% | 87%, 100.0% |
|  | False negatives | 3 (6%) | - |
| Negative | PCR | 225 | - |
|  | BD | 220 | - |
|  | Specificity (NPA) | 98% | 96%, 100% |
|  | False positives | 5 (2%) | - |
| Concordance | Cumulative | 272 | - |
|  | Agreement | 264 | - |
|  | OPA | 97% | 95%, 99% |
| Accuracy | AUC | 96% | 91%, 100% |

Abbreviations: PCR, Polymerase Chain Reaction; PPA, Positive Percent Agreement; NPA, Negative Percent Agreement; OPA, Overall Percent Agreement; AUC, Area under the curve; CI, Confidence Interval
Statistical test for categorical variables: Chi-square test

**Table 3:** Performance of BD Veritor antigen test by symptom status.

| Symptoms | Sensitivity |  |  | Specificity |  |  |
| --- | --- | --- | --- | --- | --- | --- |
|  | PCR<br>positive | BD positive (%) | 95% CI | PCR<br>negative | BD negative (%) | 95% CI |
| Asymptomatic | 19 | 16 (84%) | 68, 100.0 | 128 | 127 (99%) | 98, 100.0 |
| Symptomatic | 28 | 28 (100%) | - | 97 | 93 (96%) | 92, 100 |

Abbreviations: PCR, Polymerase Chain Reaction; CI, Confidence Interval
Statistical test for categorical variables: Chi-square test

Among the 47 specimens with positive PCR results, the mean Ct value was 16.3 **(Figure 1)**. At Ct values of between 1 and 20 (n=37), the sensitivity of the BD Veritor was 95%, while at Ct values of between 21 to 25 (n=11), sensitivity was 91%. We could not detect an association between Ct values and sensitivity of the BD veritor test at this sample size. **(Table 4)**. There was no association between Ct value and presence of symptoms (p=0.544). There was also no difference in average CT values between BD Veritor true positive and BD Veritor false negative samples (p=0.303).

**Figure 1:**
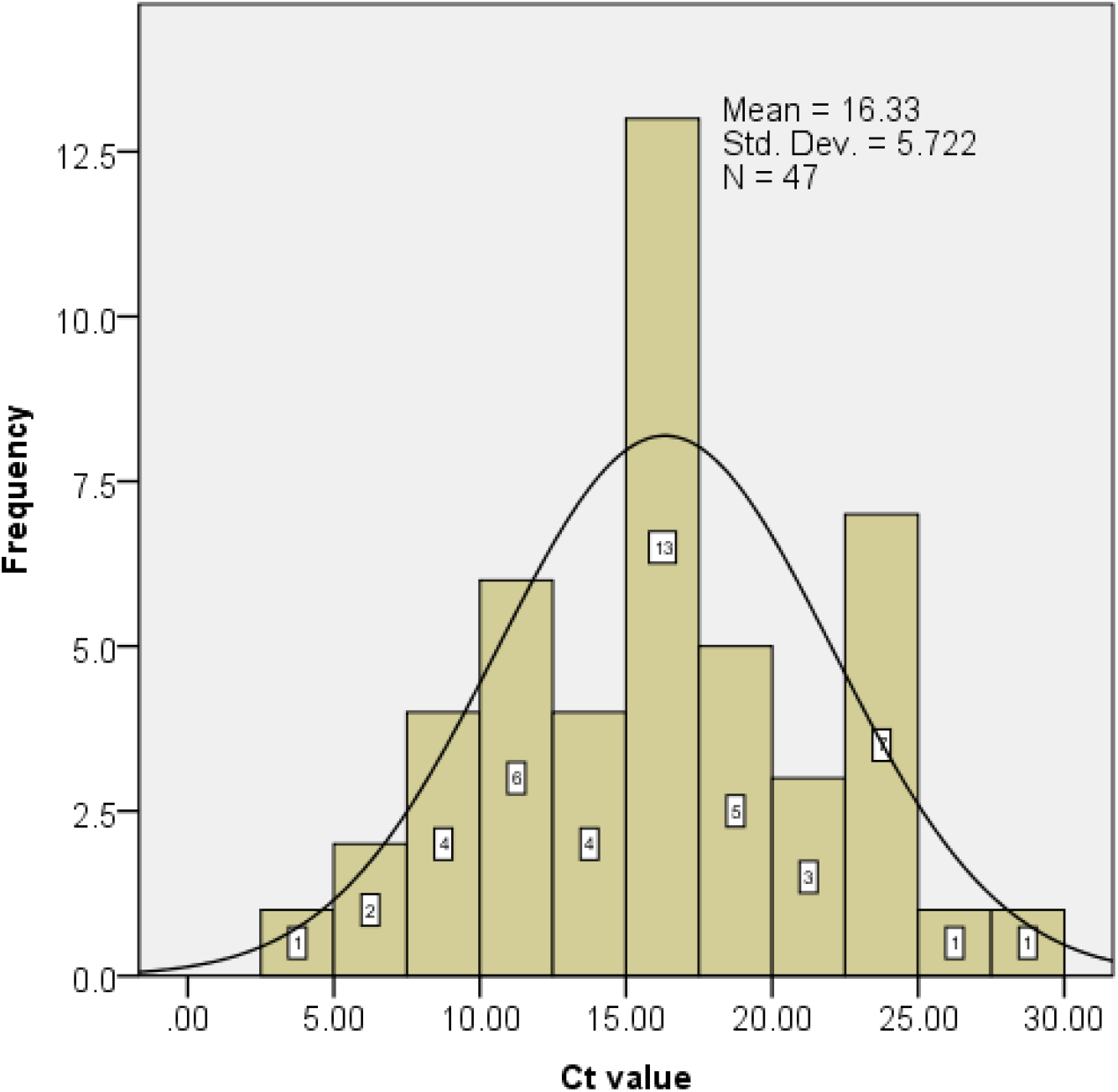
Frequency distribution of Ct values. Abbreviations: Ct, Cycle Threshold

**Table 4:** Performance of the BD Veritor test compared to Ct values.

| Ct value | Sensitivity |  |  | Specificity |  |  |
| --- | --- | --- | --- | --- | --- | --- |
|  | PCR positive | BD positive (%) | 95% CI | PCR negative | BD negative (%) | 95% CI |
| 0 to 20 | 37 | 35 (94.6%) | 87.3, 100.0 | 234 | 229 (98%) | 96.0, 99.7 |
| 21 to 25 | 11 | 10 (90.9%) | 73.9, 100.0 | 0 | - | - |
| >25 | 5 | 5 (100%) | - | 0 | - | - |

Abbreviations: PCR, Polymerase Chain Reaction; Ct, Cycle Threshold; CI, Confidence Interval
Statistical test for categorical variables: Chi-square test

There was an association observed between Ct value and length of time from specimen collection to analysis time. High Ct values were associated with longer time to analysis, with samples having a Ct value >37 (PCR negative) having a median time to analysis of 18 days, and those with Ct value of between 1 and 20 having a median time to analysis of 7 days (p<0.001).

### Implementation characteristics

#### Knowledge on testing strategies available

Clients demonstrated some level of understanding on the COVID19 testing strategies available. The key source of this information was electronic/digital media platform.

> “I have only heard and seen on TV, but I have no experience or encountered other methods.” **(KII, HCW)**
>
> “Not much, especially because Covid is a new disease and besides the normal information being shared on the media all I know is that for travelers like me, we have to take a test every time we want to travel.” **(KII, Traveler)**

Among the health care workers interviewed, there was a general indication of knowledge on current strategies in testing.

> “Yes, I am aware of the PCR gold standard testing and antibody testing.” **(KII, HCW)**.

#### Perceptions on sample collection and applicability of BD Veritor POC

Even though the clients reported the anterior nares method of sample collection as causing irritation and discomfort, it was the preferable method in comparison to oropharyngeal swabbing for RT PCR.

> “The anterior nares testing was a bit uncomfortable, but it was not painful. Comparing to the previous tests which were very invasive, this one was very friendly.” **(KII, HCW)**
>
> “Comparing to the one I had before, this test was so comfortable. Although the anterior nares sample collection was a bit uncomfortable, irritation was mild and faded away after a few minutes. This cannot be compared to the nasopharyngeal which persists for hours after sample collection” **(KII, Client)**
>
> “I wouldn’t say it was very comfortable. There was mild irritation on the nose, but it was not very invasive. I was surprised that it was not painful as earlier depicted on the TV.” (KII, Client) “Well, the test is easy and sample collection was fast and comfortable.” **(KII, HCW)**

The BD Veritor POC was preferred among traveler-participants with the main reason cited as the test’s quick turn around time in the availing of COVID19 results.

> “It was easy, and besides the sampling it took roughly 15 minutes I would prefer BD for now if it means saving time. **(KII, Traveler)**
>
> “……… I found this test quick and I think it should be approved to be used for people travelling occasionally.” **(KII Traveler)**

HCW indicated that the BD veritor POC was easy to use with going further to recommend the test for implementation in the pediatric population.

> “I would choose BD Veritor. This testing method applies to everyone especially children and patients in ICUs.” **(KII, HCW)**
>
> ……The kit application in the field is logical and easy to use and requires less training. (KII, HCW) “It was very easy, especially the nasal swab. I would prefer the BD Veritor. I believe when it will be rolled out it will be cheap.” **(KII, HCW)**

Health providers also expressed that the BD antigen test meant a reduction in the cost of care provision with cheaper COVID19 diagnosis.

> “It was very easy, especially the oral swab. I would prefer the BD Veritor. I believe when it will be rolled out it will be cheap.” **(KII, HCW)**

The short duration for COVID19 diagnosis made it largely acceptable among travelers, health providers and the public.

> “It was easy, and besides the sampling it took roughly 15 minutes. I would prefer BD for now if it means saving time. **(KII, Traveler)**
>
> “…As for retaking a test, I would prefer the BD Veritor as it gives the results much faster. I would also recommend this test to anyone willing to take a covid test based on my experience.” **(KII, Client)**

#### Participant-driven improvement suggestions

Some participants felt that there is a need for consideration of alternative methods of sample collection that minimized client discomfort. One HCW also mentioned the need for internal validation mechanisms.

> “Sample collection. I almost vomited. I wish there was another way rather than swabbing the nose.” **(KII, Traveler)**
>
> “Internal validation. The device should have the capability to print test results and have a sample counting ability. Also, I would wish if the device could display the viral load in terms of cycle number.” **(KII, HCW)**
>
> “Maybe the swab being used for the nasal sample could be made more friendly to avoid irritation.” **(KII, Client)**

## DISCUSSION

The sensitivity and specificity of BD Veritor™ System as reported by the manufacturer is 94% and 99% respectively. This study, nested in real-world use-case scenarios in Kenya, demonstrated a sensitivity and specificity of 94% and 98%, respectively, which was relatively high compared to that observed in similar studies (23) (24)(25).

Sensitivity of rapid antigen tests has been shown to be higher in symptomatic patients and in those with high viral loads. However, we did not detect an association between the presence of symptoms and sensitivity, or between the presence of symptoms and Ct value. This is potentially due to our samples size (as demonstrated by the wide confidence intervals). There was also a lack of association between Ct value and sensitivity of BD Veritor test, although we did not enroll many patients with high Ct value and were unable to adequately assess their potential effect on performance of the test.

The analysis of association between duration of symptoms and PCR results, Ct value and rapid antigen test sensitivity was carried out on a sample size of 76 participants for whom we had complete data. To assess for selection bias, we analyzed the sample characteristics of this group and compared it to that of individuals whose duration of symptoms was not recorded. There was no statistically significant difference in sex and mean age between the two groups.

When evaluating the accuracy of rapid antigen tests, factors affecting the performance of the reference standard RT-PCR must be considered. Important in this study is the source of and volume of samples taken, transport and storage conditions and the technical performance of the assay. Careful specimen collection and processing by qualified and experienced staff was carried out in order to ensure that adequate genetic material was obtained and that sample contamination was minimized. However, there were delays in analyzing test results caused by shortages of RT-PCR reagents and materials. The effect of these delays was assessed by comparing PCR results and Ct values by time from sample collection to analysis. A greater Ct value was observed in specimens for which there was an extended interval between sampling and analysis. This suggests that degradation of viral genetic material may have occurred, which may have had the effect of reducing the RT-PCR test positivity, and thus artificially increasing the sensitivity of the rapid antigen test in the study. The stringent requirements related to sample processing before analysis is a recognized drawback to RT-PCR as a testing modality in field conditions. Sample degradation is a common outcome where transport networks to central laboratories are inefficient, and where reagent stock-outs occasioned by high demand for testing are typical. This may lead to inaccuracies in reported results and missed opportunities for effective isolation and treatment to prevent forward transmission. In these conditions, point of care testing with rapid antigen tests will likely perform better than the reference RT-PCR standard. Contributing to the increased sensitivity of the rapid antigen test may also have been high viral loads (mean Ct value 16.10) in sampled participants during the peak second wave of SARS-CoV-2 infection in the country in March 2021.

Pooled testing is a strategy that is used when conducting RT-PCR assays. It is accepted as an approach that effectively identifies SARS-CoV-2 infection while conserving laboratory resources (26)(27). In addition, pooling has been shown to increase test specificity as positive samples are tested twice (28). Evidence suggests that testing accuracy is retained in pool sizes of up to 32 samples (26). In our study, samples from asymptomatic patients were included in pool sizes of 10, in line with local laboratory protocols. Deconvolution was carried out for all positive pools. The feasibility assessment in this study shed some light on the facilitators and barriers to use of rapid antigen tests. There were different levels of understanding among participants on the COVID-19 testing strategies available. Electronic platforms were the main source of information on the testing methods and the necessity for testing especially before travel. There was discomfort reported on both the anterior nares and oropharyngeal swabbing methods, but a general preference for the former. Both methods were reported to be more comfortable compared to nasopharyngeal swabbing that was most commonly depicted in the media. Most users, and especially travelers, appreciated the rapid nature of receiving results. Health care workers highlighted challenges posed by RT-PCR testing including prolonged turn-around time, high cost, equipment breakdown and rigorous sample handling requirements. They showed appreciation for the ease of use of rapid tests and postulated that they would be applicable in a wide array of settings. However, there was concern about inconclusive results and the lack of a physical report accompanying the test.

Central laboratories have as their focus the quality and reliability of a test. Clinicians and patients may in addition value methods that expedite decision making at the first point of contact(19). These include decisions to rule out life-threatening conditions, or as with COVID-19, whether or not to isolate an individual. This study demonstrated the potential for rapid antigen tests to facilitate timely clinical decision making.

Our study relied on field laboratory personnel to carry out the rapid antigen test. However, the ideal point-of-care test is one that can be used during the clinician-patient interaction. This would require additional training and mentorship of health workers in areas that are traditionally laboratory-centered, thus increasing their work-load and potentially affecting their acceptance of the method. Concerns about quality control of the tests and availability of technical support when required should be factored into any cost analysis. Also important to consider is the impact of a wrong clinical decisions resulting from inaccurate test results when employing a new method, and the effect this may have on acceptability by health care workers (19). Further studies would shed more light on these crucial aspects.

Overall, we observed an exponential increase in demand for COVID 19 testing in participating health facilities over the course of the study, indicative of a general acceptability and positive user experience with the rapid antigen test in this population.

## Conclusion

The BD Veritor rapid antigen test exhibited excellent sensitivity and specificity when used to detect SARS-CoV-2 infection among symptomatic and asymptomatic individuals in varied population settings. Its implementation feasibility, acceptability and ease-of-use in low resource settings would potentially result in bridging testing disparities between primary and tertiary health care facilities and contribute towards a reduction in community transmission. However, special protocols should be designed that distinguish workflows related to SARS-CoV-2 testing for identification and isolation of infectious individuals. Further areas of study to describe the most appropriate cadre of staff and skill-set required in busy clinical settings, as well as strategies to ensure acceptable quality of rapid antigen testing would be useful.

## Data Availability

The datasets used and/or analyzed during this study are available from the corresponding author on reasonable request.

