## Supplemental Table 1 for "ASSESSMENT OF PERFORMANCE AND IMPLEMENTATION CHARACTERISTICS OF RAPID POINT OF CARE SARS-CoV-2 ANTIGEN TESTING"

**Table 1: COVID19 symptoms among participants**

| **Variable** | **Symptomatic** | **Asymptomatic** | **P value** |
| --- | --- | --- | --- |
| **Sex (n, %)**  Female  Male | 73 (59)  52 (42) | 62 (42)  85 (58) | 0.008 |
| Mean age in years (SD) | 33 (11) | 27 (8) | <0.001 |
| **Population (n, %)**  General  HCW  OPD patient  Student  Traveler | 16 (13)  41 (33)  41 (33)  10 (8)  17 (14) | 14 (10)  34 (23)  0  80 (54)  19 (13) |  |

Abbreviations: HCW, Health Care Worker; OPD, Out-Patient Department; SD, Standard Deviation

Statistical test for categorical variables: Chi-square test; Statistical test for continuous variables: Student’s t-test
